# Unraveling Interoceptive Processing and Action Dynamics: Exploring Neural and Psychological Responses to Food Cues Using fMRI

**DOI:** 10.1101/2024.10.11.24315350

**Authors:** Nastaran Malmir, Hamed Ekhtiari, Ali Farhoudian, Somaye Robatmili, Michael Nistche

## Abstract

Interoception, the perception of body signals, which is crucial for maintaining metabolic homeostasis, is asserted to play a vital role in obesity. Despite conceptual assumptions that impaired insular interoceptive processing contributes to overeating behaviors predominantly through modulating motor cortices, this link has not been extensively explored. Therefore, to further investigate neural mechanisms underlying insula-based interoceptive processing, this pre-registered (PMC9003175) study assessed blood oxygenation level-dependent (BOLD) responses via functional magnetic resonance tomography (fMRI) in 45 healthy participants (31 females/14 males, age 35.78 ± 10 years, BMI 29.52 ± 3.5 kg/m^2^) during a block-designed food cue reactivity task. Region of interest (insula) and whole brain voxel-wise correlation analyses explored neural correlates of visceral interoception. Furthermore, group factor analysis (GFA) unveiled coherence patterns between neural (fMRI) and psychological/behavioral measures. At the psychological level, self-reported hunger (P < 0.01, d = 0.82) and food craving (P < 0.01, d = 0.68) significantly increased, while craving control (P = 0.04, d = 0.37) decreased after cue exposure. Voxel-wise correlation analysis identified positive correlations (P < 0.01) between visceral interoception and activation of the precentral gyrus (PrG or motor cortex), insula, inferior frontal gyrus (IFG), posterior cingulate cortex (PCC), and superior parietal lobule (SPL). Moreover, altered functional connectivity dynamics were noted within the insula-PrG-IFG network during food cue exposure, with a significant reduction of IFG-PrG connections (P = 0.05). Interestingly, GFA identified a cross-unit latent factor across neural and psychological/behavioral measures. Overall, our findings indicate that altered interoceptive processing (insula activity), motor planning (motor cortex activity), and diminished inhibitory control (negative IFG-PrG connectivity) collectively contribute to food craving generation and potentially subsequent actions toward food consumption.

## 1. Introduction

The act of eating is one of the most basic and earliest human behaviors and is modulated by homeostatic control mechanisms (Woods, 2009). However, the pleasurable experience of food intake may lead to overeating, raising the risk of excessive weight gain and obesity onset (Kringelbach, 2005; Stice & Burger, 2019). Despite the detrimental consequences of obesity on both psychological and physiological well-being (Ejerblad et al., 2006; Luppino et al., 2010; Mokdad et al., 2003; Park, Hong, & Park, 2017), interventions targeting weight loss and maintenance have demonstrated limited efficacy (Dombrowski, Knittle, Avenell, Araújo-Soares, & Sniehotta, 2014; Montesi et al., 2016), potentially contributing to the variability of neural and psychological responses to food cues among distinct groups of individuals with maladaptive eating behaviors (Morys, García-García, & Dagher, 2023; Murdaugh, Cox, Cook III, & Weller, 2012; Varkevisser, Van Stralen, Kroeze, Ket, & Steenhuis, 2019).

Exposure to palatable food cues predominantly triggers internal sensations of craving- an intense physiological and psychological desire toward food consumption (Jansen, 1998). Neuroimaging evidence demonstrated that viewing food images activates regions commonly referred to as the ‘appetitive network’, including the amygdala, insular cortex, precuneus, parahippocampus, postcentral gyrus, orbitofrontal cortex (OFC), and visual processing areas such as the fusiform gyrus (FuG) and lingual gyri (Tang, Fellows, Small, & Dagher, 2012). A recent meta-analysis suggested that neural responses to food cues do not differ relevantly between individuals with normal weight and obesity (Yang, Wu, & Morys, 2021).

Craving caused by food cues, termed food cue reactivity, is considered as one core neurocognitive mediator of problematic eating behaviors (Boswell & Kober, 2016; Isabel García-García et al., 2014; Jansen et al., 2003; Lawrence, Hinton, Parkinson, & Lawrence, 2012; Murdaugh et al., 2012; N Gearhardt MS, Phil, & R Corbin, 2011; Nederkoorn & Jansen, 2002; Rogers & Hill, 1989; Stice, Yokum, Blum, & Bohon, 2010; Tang et al., 2012; Winter, Yokum, Stice, Osipowicz, & Lowe, 2017). Moreover, it was proposed that subjective responses (e.g. the subjective feeling of craving or motivational state to consume food), mainly induced by external stimuli (e.g. food commercials), are initiated by changes in the body state perceived by visceral sensations (Cannon, 1987; Damasio, 1996; James, 1890). Interoception serves as a crucial component between food cue reactivity and obesity (Craig, 2002; Critchley & Harrison, 2013; Critchley, Wiens, Rotshtein, Öhman, & Dolan, 2004). It includes the ability to perceive and cognitively interpret internal body signals, and directs adaptive behaviors aimed at maintaining homeostasis in response to incoming sensory inputs (Barrett & Simmons, 2015; Craig, 2002, 2003; Seth, 2013; Wiens, 2005).

Bruch (1961) was the first to propose that inadequate interoceptive signal processing may lead to disturbed eating behaviors, supported by recent findings (Khalsa et al., 2015; Klabunde, Acheson, Boutelle, Matthews, & Kaye, 2013; Pollatos et al., 2016; Pollatos et al., 2008). This impairment encompasses an inability to accurately perceive, integrate, and translate interoceptive signals into adaptive motor responses according to the body’s homeostatic state, reflecting a vulnerability that may lead to excessive eating behavior and weight gain (Barrett & Simmons, 2015; Craig, 2009; Seth & Friston, 2016). In this study, we employed responsiveness to food cues (RFC) as a metric reflecting visceral interoceptive processing changes following food cue exposure, quantified as the difference between self-reported craving levels after food cue exposure.

Brain imaging studies have highlighted the integral role of the insula in perceiving interoceptive signals (Farb, Segal, & Anderson, 2013; Ronchi et al., 2015) and have further identified associations between obesity and insular cortex pathology, including structural abnormalities (García-García et al., 2019; Herrmann, Tesar, Beier, Berg, & Warrings, 2019; Janowitz et al., 2015; Saute, Soder, Alves Filho, Baldisserotto, & Franco, 2018), as well as altered connectivity patterns (Avery et al., 2017), and dysfunctional activity (Avery et al., 2015; Contreras-Rodríguez et al., 2019; Simmons et al., 2013; Uddin, 2015).

As a major node within the interoceptive system, the insula is characterized by its involvement in a wide range of sensory, emotional, and cognitive processes (Seeley et al., 2007; Uddin, Kinnison, Pessoa, & Anderson, 2014). Moreover, the insula has a multi-layered structure that directs the flow of interoceptive information from more posterior granular to more anterior agranular regions (Craig, 2002, 2009; Kurth, Zilles, Fox, Laird, & Eickhoff, 2010; Simmons et al., 2013). In this way, sensory information is postulated to undergo initial processing in the mid-and posterior insula before being conveyed to the anterior insula for high-level cognitive processing, wherein sensory inputs are supposed to be integrated into emotional, cognitive, and motivational data derived from a complex array of cortical and subcortical regions (Craig, 2002). While food cue reactivity studies have frequently explored associations between altered interoceptive processing and aberrant activity within the mid-to posterior insula (Avery et al., 2015; Avery et al., 2017; Simmons et al., 2013; Stice & Burger, 2019; van der Laan, De Ridder, Viergever, & Smeets, 2011), other studies, including a recent meta-analysis, have focused on the crucial role of the anterior insula in processes related to cognitive control (Chang, Yarkoni, Khaw, & Sanfey, 2013; Molnar-Szakacs & Uddin, 2022; Nord, Lawson, & Dalgleish, 2021; Uddin, 2015). More specifically, Sridharan et al. (2008) highlighted the important role of the anterior insula within the saliency network (SN), emphasizing its extensive connections with key nodes of both, the default mode network (DMN) and the central executive network (CEN) (Sridharan, Levitin, & Menon, 2008), enabling coordination of respective network dynamics for optimal executive control in response to a changing external environment (Huang et al., 2021; Margulies & Uddin, 2019; Uddin, 2015). Anterior insula activation may lead to the deactivation of the DMN, impacting core regions such as the posterior cingulate cortex (PCC), and ventromedial prefrontal cortex (vmPFC), while simultaneously activating the CEN, promoting adaptive responses to salient events (Sridharan et al., 2008). Nevertheless, despite the significant emphasis put on the role of the insular cortex in processing interoceptive signals in the obesity literature (Avery et al., 2015; Barrett & Simmons, 2015; Contreras-Rodríguez et al., 2019; Molnar-Szakacs & Uddin, 2022; Rapuano, Huckins, Sargent, Heatherton, & Kelley, 2016; Simmons et al., 2013), the insula and its subregions remain among the least comprehended areas in current neuroimaging research (Menon et al., 2020). Besides, previous meta-analyses have shown that exposure to food cues and subsequent interoceptive sensations can induce actions toward food consumption (Boswell & Kober, 2016; Chen & Zeffiro, 2020; N Gearhardt MS et al., 2011; Tang et al., 2012), it is also evident that not all overweight/obese individuals exposed to food cues exhibit the same overeating behaviors (Giuliani & Berkman, 2015; Giuliani, Merchant, Cosme, & Berkman, 2018; Volkow & Wise, 2005).

Therefore, by meticulously exploring insular interoceptive processing, as the critical link between food cue reactivity and subsequent food intake, this research aims to advance the field by providing deeper insight into the neural and psychological components underlying overeating behaviors, thereby contributing to the development of more effective interventions that address the complexities of obesity. To this purpose, we designed a functional magnetic resonance tomography (fMRI) study to unravel the neural and psychological/behavioral correlates of visceral interoception, modulated by the food cue reactivity paradigm, in overweight/obese individuals. Initially, we explored brain activation linked to interoceptive processing following exposure to food cues, investigating whether respective activation patterns can be traced back to subjective responses to food cues. Importantly, building on extensive prior research in the field of insular interoceptive processing (Avery et al., 2015; Avery et al., 2017; Barrett & Simmons, 2015; Chang et al., 2013; Contreras-Rodríguez et al., 2019; Craig, 2009; Kurth et al., 2010; Molnar-Szakacs & Uddin, 2022; Nord et al., 2021; Simmons et al., 2013; Sridharan et al., 2008; Stice & Burger, 2019; Uddin, 2015; van der Laan et al., 2011), we selected the insula as a main region of interest (ROI). The insula comprises several subregions, each associated with distinct functions involved in a wide variety of sensory, emotional, and cognitive processing (Seeley et al., 2007; Uddin et al., 2014; X. Wang, Tan, Van den Bergh, von Leupoldt, & Qiu, 2020; Yarkoni, Poldrack, Nichols, Van Essen, & Wager, 2011). In the present study, we investigated all subregions of the insular cortex identified in the brainnetome atlas, comprising six bilateral ROIs, based on their known involvement in interoceptive processing (Avery et al., 2015; Avery et al., 2017; Kerr et al., 2016; Simmons et al., 2013). Moreover, we conducted a voxel-wise correlation analysis to identify brain regions functionally associated with RFC ratings independent from the predefined ROI. Specifically, in this connection we conducted an exploratory seed-to-seed functional connectivity analysis focusing on regions showing positive correlations with RFC scores. This allowed us to delve into the potential functional connectivity patterns related to the food cue reactivity task in individuals with heightened RFC ratings. Finally, we employed a group factor analysis (GFA) to explore coherence patterns between neural and psychological/behavioral measures. We hypothesized that the insula is a critical structure for interoceptive processing resulting in the subjective feeling of craving. Additionally, we predicted that enhanced insular activity would be accompanied by increased activity of motor cortices, reflecting motor readiness for food consumption. Overall, we aimed to explore how brain regions and networks are involved in the modulation of RFC ratings following exposure to food cues in overweight/obese individuals.

## 2. Method

### 2.1. Participants

Participants were recruited through nutrition clinics, flyers, and online advertisements. All potential participants were prescreened for primary eligibility criteria including age, BMI, frequent food craving, and lack of serious medical/neurological illness by an initial telephone interview. A total of 52 potential candidates of both sexes were scheduled for an initial in-person session, where they underwent baseline assessments, received a detailed explanation of the study procedure, signed the informed consent form, and became familiar with the food cue reactivity task.

To be eligible for the current study, participants were required to have a BMI greater than 25 kg/m^2^ (within the range of 25-35 kg/m^2^), be between 18 and 60 years old, and report frequent food cravings (≥3 per day during the last month), as assessed by a self-report questionnaire.

Participants were excluded if they met serious medical/mental health conditions, reported in the demographic questionnaire evaluated by a clinician, fulfilled the criteria for alcohol and substance use disorder (diagnostic and statistical manual of mental disorders, 5^th^ edition (DSM-5, or eating disorder, assessed by the eating disorder diagnostic scale (EDDS). Further exclusion criteria were a history of claustrophobia, MRI contraindications, and participation in any ongoing diet program or professional sport. Eligibility was determined by trained study staff during screening and baseline assessments.

This study was conducted at the National Brain Mapping Lab in Tehran, Iran. All participants provided informed consent according to the study protocol approved by the ethics committee of research, Iran University of Medical Sciences (IR.IUMS.REC.1396.0459).

### 2.2. Study Design

The study design and methodology presented in this paper are integral components of a larger research protocol (ID: PMC9003175, https://pubmed.ncbi.nlm.nih.gov/35413923/). **Figure 1** outlines the study course. Those participants who were identified as eligible for preliminary screening underwent two stages of the experiment on separate days. During the first visit, participants completed the baseline assessment including the gathering of demographic data, and clinical assessments, including the depression anxiety stress scales-21 (DASS-21), the eating disorder diagnostic scale (EDDS), the compulsive eating scale (CES), three-factor eating questionnaire (TFEQ), food craving questionnaire-trait (FCQ-T), and food craving questionnaire-state (FCQ-S).

**Figure 1.**
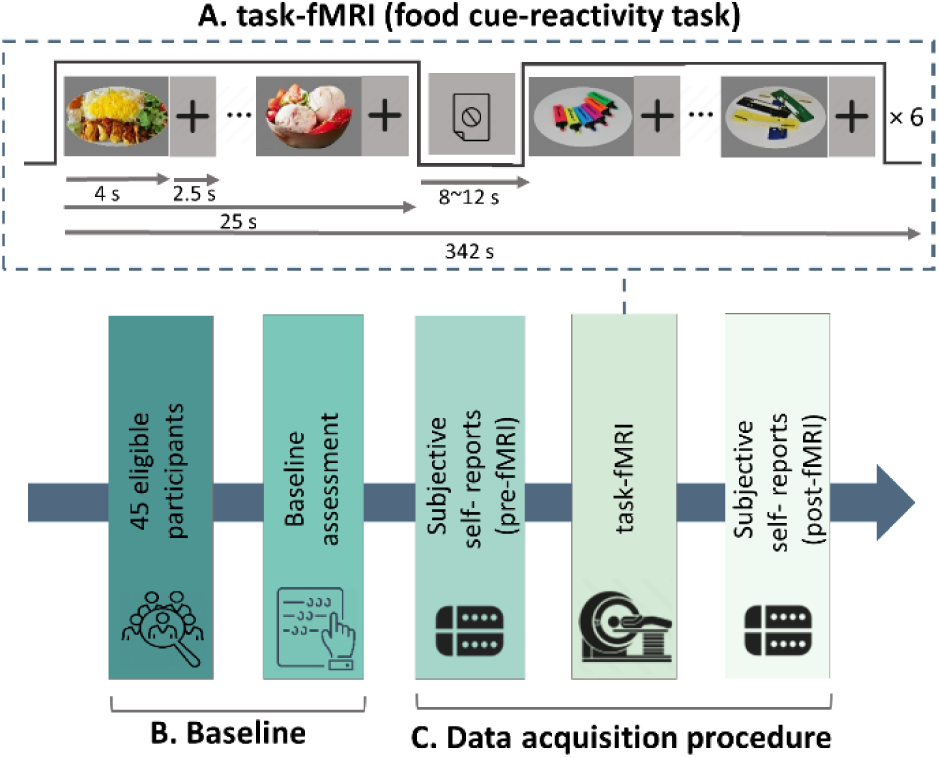
Diagram of the study design and data acquisition: **(A)** The cue-induced food craving task followed a block design, each consisting of 6 images of either food or neutral cues presented for 4 seconds, with an inter-block time interval of 8-12 seconds and an interstimulus interval of 2.5 seconds. In total, the task comprised a total of 72 cues and lasted for 342 seconds. **(B)** Forty-five eligible participants first underwent baseline assessments, including self-reports and evaluations of clinical and eating-related traits. **(C)** Each participant completed subjective assessments (self-report of food craving, hunger, craving control, and affective status) pre- and post-imaging, which included T1-weighted structural and task-fMRI scans.

This was followed by an orientation session involving answering food craving and hunger-related questions on a visual analogue scale (VAS), selecting a number from 0 (not at all) to 100 (extremely), immediately before and after the fMRI scan **(Figure 1-C)**. Food craving was operationally defined as an urge to consume food and measured by these two questions: (1) “What is your current food craving?” and (2) “How much do you feel the need to eat right now?”. The subjective feeling of hunger was rated by the following question “Please specify your current hunger” and control over craving was assessed with the question “How much control do you feel about eating right now?”. Additionally, participants were asked to rate their affective status in six domains—happiness, sadness, anxiety, anger, drowsiness, and awareness— on a VAS scale ranging from 0 to 100. For example, they were asked, “Please rate your current feeling of happiness”.

Following the completion of the scanning session, participants were again asked to rate their craving, hunger, craving control, and affective status on the VAS scales. All assessments and stages of the experiment were conducted by well-trained researchers.

### 2.3. Food Cue Reactivity Task

The food cue reactivity task was conducted while the participants underwent MRI, as illustrated in **Figure 1-A**. The task had a block design comprising 12 blocks, with 6 blocks featuring food images and 6 blocks neutral images. Each block consisted of 6 images displayed for 4000 ms each in randomized order, followed by a jittered inter-block interval lasting between 8 and 12 seconds. The experimental paradigm thus comprised a total of 72 cues, lasting 342 seconds. All images were presented on a gray background at the center of an LCD screen located in the scanner, directly in front of the participant’s face (the eye-monitor distance was approximately 20cm). Participants were instructed to attentively view the cues without distraction. Food images were sourced from the Internet and reviewed by two independent assessors. Neutral images originated from the Full4Health Image Collection (Charbonnier, van Meer, van der Laan, Viergever, & Smeets, 2016), with content and quality matched to the food images **(Figure 1-A)**.

### 2.4. MRI Protocol and Data Acquisition

Subjects were equipped with earplugs and headphones for noise protection during the scan, and head motions were restricted using an MRI head positioning pad placed around the head. Participants were explicitly instructed that even small movements could impair imaging quality. Neuroimaging data were acquired on a 3 Tesla scanner (MAGNETOM Prisma, SIEMENS, Germany) with a 20-channel head coil at the National Brain Mapping Lab, Tehran, Iran. The MRI session comprised a standard T1-weighted MPRAGE (magnetization-prepared rapid-acquisition gradient echo) scan, followed by a task-based functional MRI using the food cue reactivity paradigm **(Figure 1)**. Structural T1-weighed images were acquired with the following parameters: TR = 1810 ms, TE = 3.45 ms, TI = 1100 ms, 176 slices, 1.03 × 1.03 × 1.0 mm3, flip angle = 7°. Task-based fMRI parameters included: TR = 2500 ms, TE = 23 ms, FOV = 192 × 192 mm2, 43 slices, 167 volumes, 3 × 3 × 3 mm3, flip angle FA = 7°.

### 2.5. Food Cue Reactivity Task fMRI Analysis

The task-based functional MRI data underwent preprocessing and analysis using FSL v6.0.4, Functional MRI of the Brain (FMRIB) analysis group software library, Oxford, UK (Jenkinson, Beckmann, Behrens, Woolrich, & Smith, 2012).

The preprocessing steps included motion correction using the MCFLIRT toolbox, skull-stripping with the brain extraction tool, and slice timing correction based on interleaved slice acquisition differences. Subsequently, functional images were spatially normalized to echo-planar images (EPI) templates constructed in standard Montreal neurological institute (MNI) space and smoothened with a 5 mm full-width at half-maximum Gaussian kernel.

Following preprocessing, we employed the FEAT toolbox, FSL, based on a general linear model (GLM) statistical analysis combined with a standard hemodynamic response function (HRF), to conduct a whole brain subject-level analysis of the main contrast of interest (food > neutral cues). The individual contrast maps were then combined in a group-level analysis to examine the mean neural response to the main condition of interest (food > neutral cues). Group-level analysis was performed via FMRIB’s local analysis of mixed effects (FLAME), and significant clusters were identified with a voxel-level probability threshold of Z > 3.1, corrected for multiple comparisons by false discovery rate (FDR) correction at P < 0.05 (Worsley et al., 1996).

### 2.6. Region of Interest Approach

Based on the aforementioned interoception literature, we primarily focused on the insula as the main region of interest (ROI). Bilateral masks were created using the featquery tool from FSL (FEAT, www.fmrib.ox.ac.uk/fsl) for all subregions of the insula, and anatomically and functionally defined based on the Brainnetome atlas (Fan et al., 2016), including the hypergranular (G), ventral agranular (vIa), dorsal agranular (dIa), ventral dysgranular and granular (vId/vIg), dorsal granular (dIg), and dorsal dysgranular (dId) divisions, totaling in 12 subregions.

In addition, to determine whether responsiveness to food cue (RFC) ratings − a measure indicative of visceral interoception, computed as the disparity between post-fMRI craving and pre-fMRI craving − was associated with specific regional brain activities during the food cue reactivity task, we conducted a whole brain voxel-wise correlation analysis for the contrast food > neutral by adding RFC scores as a covariate of interest. For each participant, RFC scores were included as a covariate to the GLM in the group-level fMRI analysis. Z statistic images were thresholded using clusters determined by Z > 2.3 and clusters with P < 0.05 corrected for multiple comparisons using FDR were considered significant.

Regarding the observed substantial activation of the precentral gyrus (PrG) (P < 0.001 and k = 1176 voxels) in individuals who reported higher ratings of RFC, along with evidence suggesting a potential relationship between insular interoceptive processing and motor readiness for food consumption (see Introduction and Discussion sections for more details), we selected the PrG as the second ROI and examined the blood oxygenation level-dependent (BOLD) responses across its six bilateral subregions during the food cue reactivity task, including the head and face, caudal dorsolateral, upper limb, trunk, tongue and larynx, and caudal ventrolateral areas in a total of 12 subregions. For each participant, we extracted mean parameter estimates of the BOLD signal strength induced by food cue exposure from each of the 24 ROIs (12 subregions of the insula and 12 of the PrG) in the Brainnetome atlas, derived from the subject-level analyses for the main contrast of interest (food > neutral).

### 2.7. Exploratory Task-based Functional Connectivity Analysis

Task-based functional connectivity analysis was performed using generalized psychophysiological interaction (PPI) analysis implemented in the CONN toolbox (version 20.b) (Whitfield-Gabrieli & Nieto-Castanon, 2012).

In this analysis, we employed a hypothesis-driven, seed-to-seed functional connectivity approach to assess functional connectivity between brain regions showing positive correlations with RFC ratings in response to food cues (contrast: food > neutral) (refer to **Figure 4-B** and **Supplementary Table 2** for detailed information). Therefore, the chosen seed regions included the insula (coordinates: −30, 18, 8), PrG (coordinates: −32, −8, 54), inferior frontal gyrus (IFG) (coordinates: 54, 16, 14), caudodorsal cingulate (coordinates: 0, 18, 34), and superior parietal lobule (SPL) (coordinates: 22, −52, 54). We explored if these clusters showed significant functional connectivity with each other in response to food cues compared to neutral cues. For the PPI first-level analysis, three regressors (timings of both food and neutral blocks, time series of the predefined seed regions, and the PPI term) were created for each participant, and only significant regional connectivities surviving FDR correction at a cluster-level threshold of P < 0.05 are reported.

### 2.8. Group Factor Analysis

Group factor analysis (GFA) is a Bayesian analysis technique employed to describe relationships between different sets of variables by identifying latent factors across multiple datasets (Klami, Virtanen, Leppäaho, & Kaski, 2014). Here we applied GFA to detect a relationship between three groups of variables, namely neural, psychological/behavioral, and demographic measures.

#### 1. Neural measures (28 variables)

This block included brain regions showing positive subjective-neural correlations during the food cue reactivity task, as identified by voxel-wise whole brain analysis, which included the bilateral insula *(hypergranular, ventral agranular, dorsal agranular, ventral dysgranular and granular, dorsal granular, and dorsal dysgranular areas)*, bilateral PrG *(head and face, caudal dorsolateral, upper limb, trunk, tongue and larynx, and caudal ventrolateral areas)*, IFG, caudodorsal cingulate, and SPL.

#### 2. Psychological/behavioral measures (18 variables)

This block included the following assessments: self-reports of baseline craving, as well as changes in craving (or RFC), hunger, and craving control after food cue exposure, as rated on a VAS ranging from 0 to 100. Additionally, participants completed food-related questionnaires during baseline assessment, including the CES, TFEQ *(subscales: cognitive restraint of eating, emotional eating, and uncontrolled eating)*, FCQ-T *(subscales: lack of control, preoccupation, hedonic hunger, emotions, and guilt)*, and FCQ-S *(subscales: intense desire, anticipation of rewarding, anticipation of relief, lack of control, craving as a physiological state)*.

#### 3. Demographic Measures (3 variables)

This block included BMI, age, and education. Collectively, the GFA model comprised a total of 49 variables and was executed using R package GFA 4.0.5. We adapted the variables for GFA by z-normalizing them to achieve a zero mean and unit variance. To enhance the reliability of latent factor identification and minimize the risk of erroneous results, we conducted the GFA estimation process 10 times, maintaining consistent robust latent factors across sample chains.

### 2.9. Further statistical Analyses

Subjective self-reports, psychometric questionnaires, and demographic characteristics were descriptively analyzed by the Statistical Package for the Social Sciences (SPSS. version 24). Mean differences in self-reported food craving, hunger, craving control, and affective status before and after imaging were compared using paired sample t-tests (significance level: P < 0.05, with Cohen’s d indicating effect size). Additionally, Pearson correlation analyses were conducted to examine the correlation between RFC scores and mean activation values within each subregion of the two main ROIs, comprising a total of 24 ROIs− 12 within the insula and 12 within the PrG.

## 3. Results

In this study, out of 52 potential candidates, two opted to withdraw from the study after the baseline session and one was not eligible due to diagnostic criteria for severe obsessive-compulsive disorder. Additionally, one participant had to be excluded due to unanticipated claustrophobia, another for an intracranial lesion, and two for poor data quality, so their data were excluded from further analysis. The final sample of 45 subjects (31 females, 14 males, age = 35.78 ± 10 years, ranging from 20 to 55 years, BMI = 29.52 ± 3.5 kg/m2, range 24-40) were included in the further assessment. Detailed demographic variables, clinical characteristics, and eating-related statistics can be derived from **Table 1**.

**Table 1.**
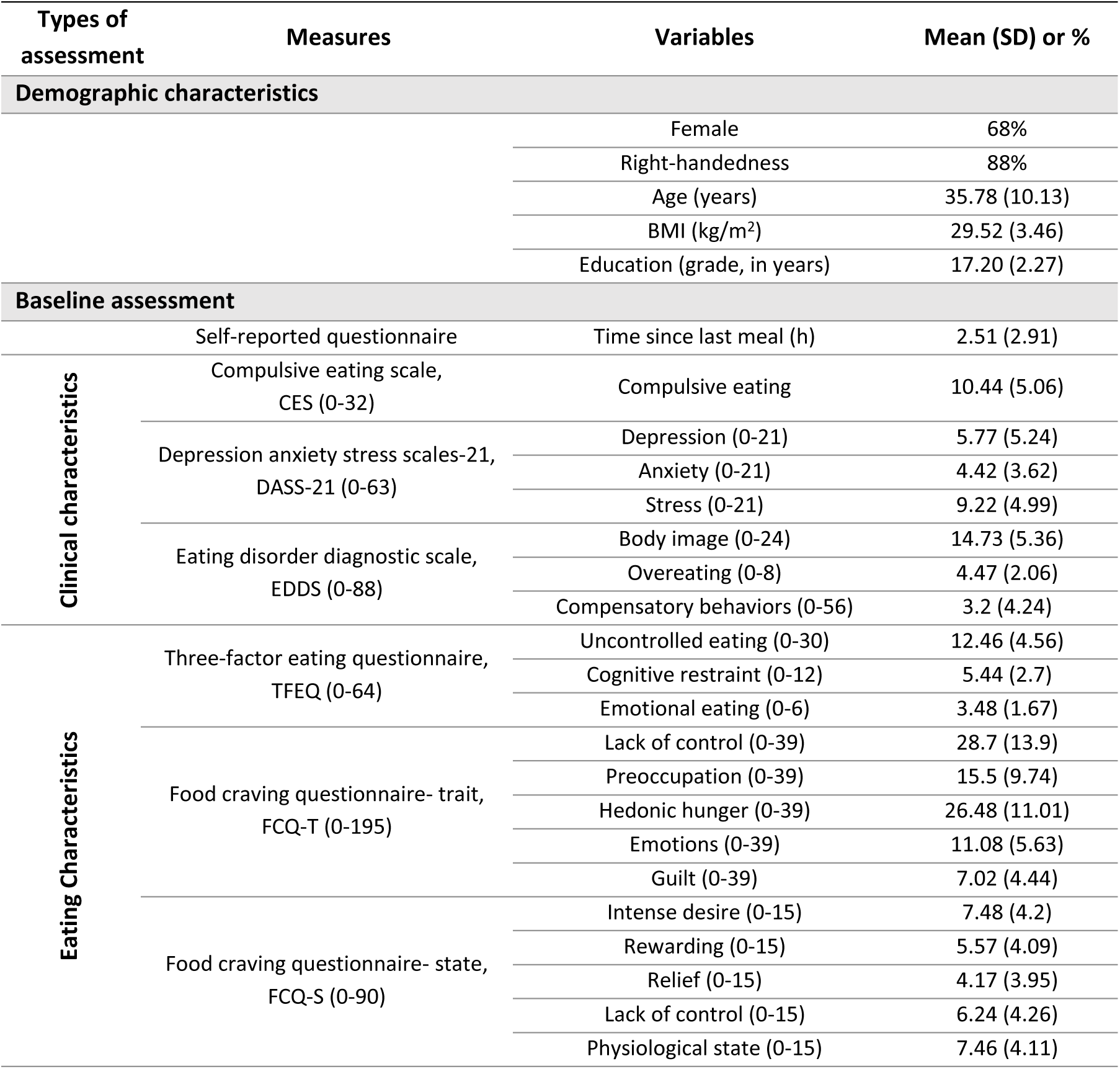
Descriptive statistics of demographic characteristics and baseline measures across all overweight /obese participants (n=45).

### 3.1. Baseline Data

Forty-five overweight/obese participants (68% female) with a mean BMI of 29.52 ± 3.46 kg/m2 were included in the analysis. Descriptive statistics for demographic, clinical, and eating characteristics are presented in **Table 1**.

Participants, on average, reported 2.5 hours since their last meal (SD = 2.9) as planned, to standardize the time since the last meal and maintain a more neutral hunger state (36 ± 26.4) and food craving (34.33 ± 28.01) level. None of the clinical measures fell within the pathologic range.

### 3.2. Psychological/behavioral Data

The results of the 2-tailed paired sample t-test indicated a significantly increased food craving (t = 4.56, P < 0.001, d = 0.68) as well as hunger level (t = 4.49, P < 0.001, d = 0.82) after food cue exposure, as rated on a 0–100 VAS scale. Conversely, craving control decreased at the end of the experiment (t = −2.09, P = 0.04, d = −0.37). Based on Cohen’s d estimation (Kim, 2015), the effect size for hunger level and craving (referred to as responsiveness to food cues, RFC) was large, while a negative effect with a ‘medium’ effect size for craving control was identified.

In addition, affective status VAS (0-100) ratings showed increased subjective feelings of happiness (t = 2.2, P < 0.03, d = 0.33), and decreased anxiety (t = −3.37, P = 0.01, d = −0.5) and awareness (t = −2.89, P = 0.01, d = −0.43), while other affects did not significantly differ before and after food cue exposure **(Table 2)**. Comparisons of food craving before and after exposure to the food cue reactivity task and the distribution of RFC scores across all participants are shown in **Figures 2A** and **B**.

**Table 2.** Changes in craving and affective status after food cue exposure. Results of the paired t-tests for the self-reports across the full sample (n=45).

| Variables | Pre-fMRI | Post-fMRI | Paired Differences | t | P | d |
| --- | --- | --- | --- | --- | --- | --- |
| **Statistics related to food craving (0-100)** |  |  |  |  |  |  |
| Craving (RFC) | 34.33 (28.01) | 51.88 (32.39) | 17.56 (25.82) | 4.56 | <.001 | 0.68 |
| Hunger | 35.67 (27.62) | 52 (29.29) | 16.33 (19.91) | 4.49 | <.001 | 0.82 |
| Craving control | 69.06 (25.57) | 57.19 (28.31) | -11.88 (32.07) | -2.09 | 0.04 | -0.37 |
| **Affective states (0-100)** |  |  |  |  |  |  |
| Happiness | 61.11 (22.2) | 65.33 (23.51) | 4.22 (12.88) | 2.2 | 0.03 | 0.33 |
| Anxiety | 27.78 (23.73) | 17.11 (16.46) | -10.67 (21.26) | -3.37 | 0.01 | -0.50 |
| Drowsiness | 40.76 (26.01) | 35.78 (27.59) | -4.98 (32.56) | -1.03 |  |  |
| Sadness | 16 (18.75) | 13.78 (17.61) | -2.22 (14.28) | -1.04 |  |  |
| Anger | 7.33 (11.56) | 4.89 (9.44) | -2.44 (10.26) | -1.60 |  |  |
| Awareness | 88.67 (16.45) | 82.89 (20.62) | -5.78 (13.40) | -2.89 | 0.01 | -0.43 |

Results are presented as mean (SD).

**Figure 2.**
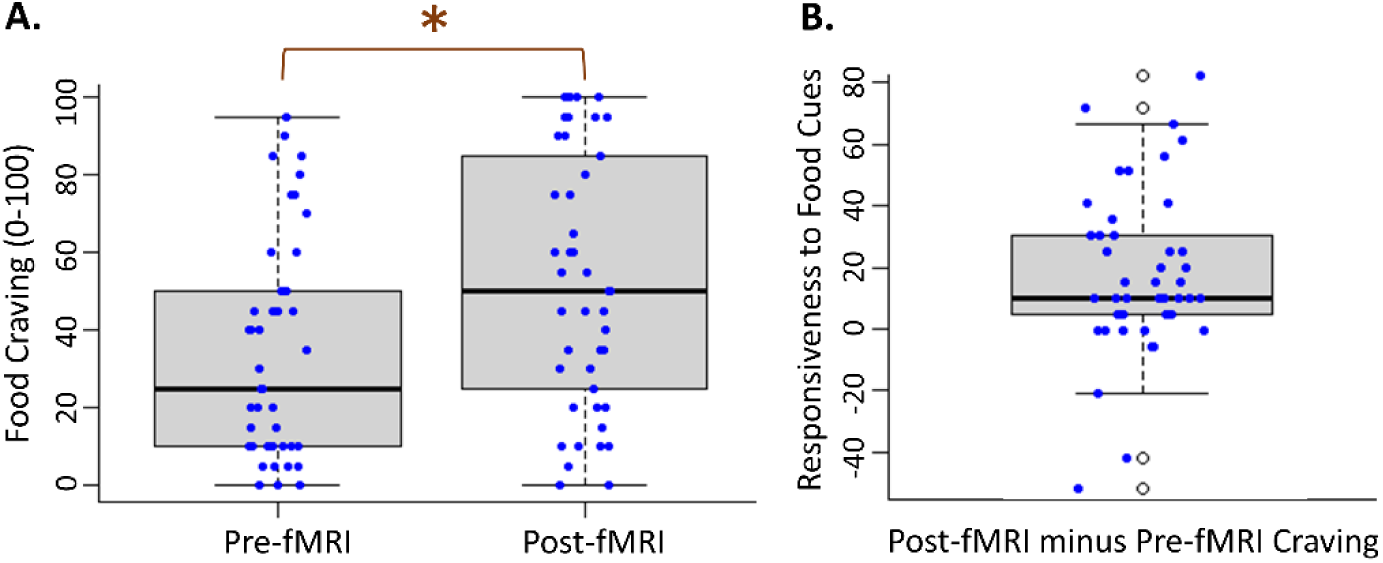
Subjective response to food cue exposure. Scatter boxplots represent **(A)** Food craving scores before and after the food cue reactivity task. **(B)** Distributions of responsiveness to food cues scores (post-fMRI food craving minus pre-fMRI food craving) across participants. In each scatter boxplot, blue circles represent the individual data and the central black line indicates the median of the data. * P < 0.01

### 3.3. Neuroimaging Data

#### 3.3.1. Group analysis in contrast food > neutral cues

As illustrated in **Figure 3**, the whole brain analysis revealed significant activation clusters during the food cue reactivity task for the contrast food > neutral in the right fusiform gyrus (FuG), left amygdala, right posterior cingulate cortex (PCC), and left superior frontal gyrus (SFG). For detailed information about the activated regions, please refer to **Supplementary Table 1**. Significant clusters were determined using the voxel-level statistical threshold of Z > 3.1, providing greater assurance of identifying activated clusters. We present all functional MRI results at the whole brain level, correcting for multiple comparisons using the false discovery rate (FDR) at a cluster threshold of P < 0.05, and determined coordinates according to the Brainnetome atlas.

**Figure 3.**
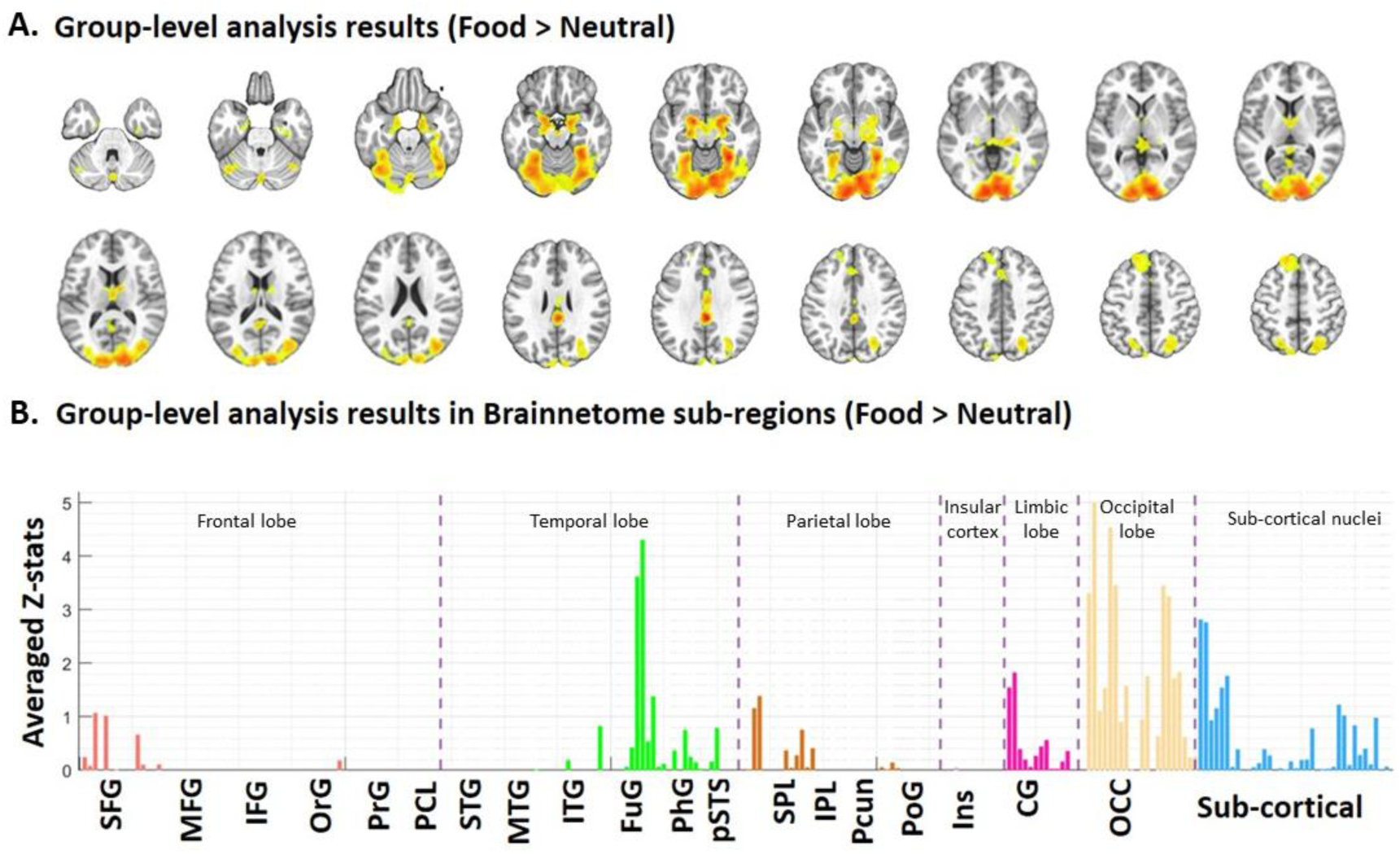
Whole brain response to the food cue reactivity task for the of food > neutral contrast. **(A)** Results of the whole brain analysis displayed greater neural activity in response to food cues relative to nonfood cues (n=45) in the fusiform gyrus, medial amygdala, posterior cingulate cortex, and superior frontal gyrus (Supplementary Table 1 for numerical results). **(B)** Brain activations are presented in the Brainnetome atlas. Z-statistic maps are whole brain clusters corrected with Z > 3.1 and P_FDR_ < 0.05. ***Note.*** *SFG* superior frontal gyrus, *MFG* middle frontal gyrus, *IFG* inferior frontal gyrus, *OrG* orbital gyrus, *PrG* precentral gyrus, *PCL* paracentral lobule, *STG* superior temporal gyrus, *MTG* middle temporal gyrus, *ITG* inferior temporal gyrus, *FuG* fusiform gyrus, *PhG* parahippocampal gyrus, *pSTS* posterior superior temporal sulcus, *SPL* superior parietal lobule, *IPL* inferior parietal lobule, *Pcun* precuneus, *PoG* postcentral gyrus, *Ins* insular gyrus, *CG* cingulate gyrus and *OCC* occipital lobe.

#### 3.3.2. Voxel-wise correlation analysis

We conducted an exploratory whole brain voxel-wise correlation analysis to identify additional functional correlates of RFC ratings in overweight/obese participants exposed to the food cue reactivity task (P < 0.05, FDR corrected, Z threshold > 2.3).

Notably, the left precentral gyrus (PrG) (k = 1176, P_FDR_ < 0.001, Z = 4.05) and left insular gyrus (k = 738, P_FDR_ < 0.001, Z = 4.33) exhibited the strongest positive correlations with increased RFC scores. Additionally, the right inferior frontal gyrus (IFG) (k = 544, P_FDR_ = 0.007, Z = 3.92), right caudodorsal cingulate (k = 522, P_FDR_ = 0.009, Z = 3.43), and right superior parietal lobule (SPL) (k = 470, P_FDR_ = 0.01, Z = 4.14) showed positive correlations with RFC scores **(Figure 4)**. **Supplementary Table 2** provides cluster-level statistics.

**Figure 4.**
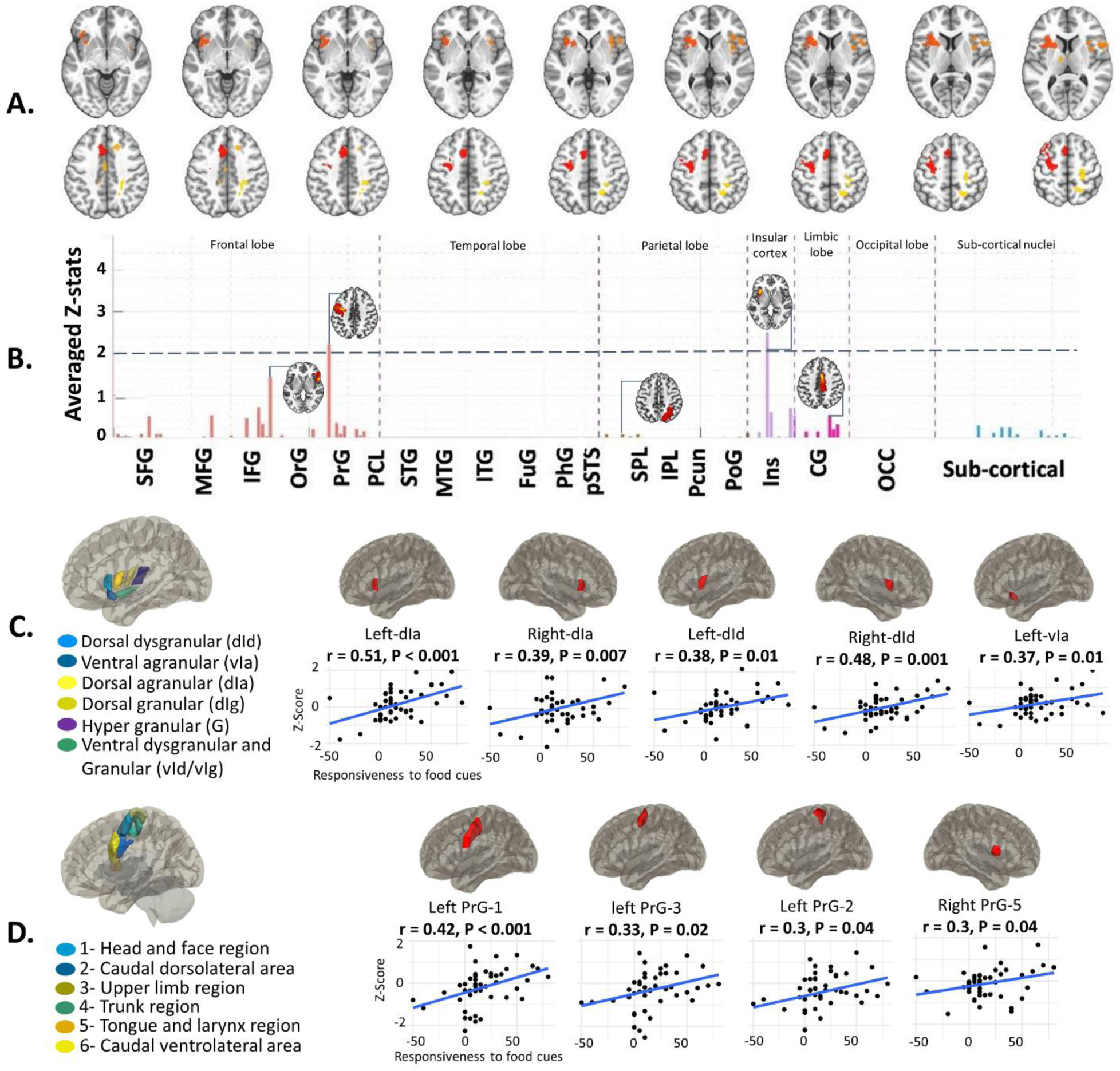
Associations between the subjective response to food cues and brain activation. Responsiveness to food cue (RFC) scores, computed as the difference between post-fMRI craving and pre-fMRI craving, was modeled as a covariate in the group-level fMRI analysis. **(A)** Voxel-wise correlation between RFC scores measured on the VAS scale (ranging from 0-100) and BOLD response to the food > neutral contrast in the full sample (n = 45) (Z > 2.3, corrected, P_FDR_ = 0.05). **(B)** Brain regions defined by the Brainnetome atlas with significant positive voxel-wise correlations. Higher RFC scores were positively correlated with BOLD activation in the PrG, insula, IFG, PCC, and SPL (see Supplementary Table 2 for numeric results). **(C)** Results of ROI-based analysis and scattering diagrams of the insular subregions showing significant correlations with RFC ratings (mid-to anterior insula including dIa, dId, and vIa). **(D)** Results of the ROI-based analysis and associated scattering diagrams for the PrG subregions with significant correlations with RFC ratings (head and upper limb areas). ***Note.*** *SFG* superior frontal gyrus, *MFG* middle frontal gyrus, *IFG* inferior frontal gyrus, *OrG* orbital gyrus, *PrG* precentral gyrus, *PCL* paracentral lobule, *STG* superior temporal gyrus, *MTG* middle temporal gyrus, *ITG* inferior temporal gyrus, *FuG* fusiform gyrus, *PhG* parahippocampal gyrus, *pSTS* posterior superior temporal sulcus, *SPL* superior parietal lobule, *IPL* inferior parietal lobule, *Pcun* precuneus, *PoG* postcentral gyrus, *Ins* insular gyrus, *CG* cingulate gyrus and *OCC* occipital lobe. *INS-G* hypergranular insula, *INS-vIa* ventral agranular insula, *INS-dIa* dorsal agranular insula, *INS-vId/vIg* ventral dysgranular and granular insula, *dIg* dorsal granular insula, *dId* dorsal dysgranular insula.

#### 3.3.3. Region of Interest Analysis

With the a priori focus on examining the role of the insula in visceral interoception via its activity modulation by the food cue reactivity task, we investigated mean parameter estimates of the blood oxygenation level-dependent (BOLD) signal intensity (contrast: food > neutral) within each subregion of the insula in the human Brainnetome atlas and then calculated correlations of these areas with RFC ratings **(Figure 4-C)**.

The correlations of the BOLD response contrast of food and neutral cues, and individual RFC ratings within the bilateral dorsal agranular (left; r = 0.51, P_FDR_ < 0.001 and right; r = 0.39, P_FDR_ = 0.007), bilateral dorsal dysgranular (left; r = 0.38, P_FDR_ = 0.01 and right; r = 0.48, P_FDR_ < 0.001), as well as the left ventral agranular area (r = 0.37, P_FDR_ = 0.01) were significant. Significant correlations were not observed for other insular subregions. **Supplementary Figure 1** provides scatter plots illustrating the correlation between RFC scores and activation of all insular subregions.

Furthermore, considering that the PrG was functionally associated with RFC measures in our whole brain voxel-wise correlation analysis (significant at P_FDR_ < 0.01 and k= 1176, k indicates the number of voxels), we proceed to conduct an additional ROI analysis to explore the parameter estimates of the signal change within all subregions of the PrG in the Brainnetome atlas. **Figure 4-D** shows PrG subregions with a significant correlation with RFC ratings. Higher RFC scores were significantly correlated with enhanced BOLD activation within the head and face (r = 0.42, P_FDR_ < 0.001), upper limb (r = 0.33, P_FDR_ = 0.02), caudal dorsolateral (r = 0.3, P_FDR_ = 0.04), and tongue and larynx (r = 0.3, P_FDR_ = 0.04) areas of the PrG in response to food cues relative to neutral cues. **Supplementary Figure 2** provides scatterplots illustrating the correlation between RFC ratings and the neural response of all PrG subregions to food cues.

#### 3.3.4. Task-based functional connectivity analysis

The generalized psychophysiological interaction (PPI) analysis revealed decreased connectivity during exposure to food cues between the caudal dorsolateral PrG-L (coordinates: −32, −8, 54) and ventral IFG-R (coordinates: 54, 16, 14) (T = −2.59, P_FDR_ = 0.05) for the main contrast of interest, food > neutral cues. Similarly, the caudodorsal CG-R (coordinates: 0, 18, 34) showed diminished connectivity with the rostral SPL-R (coordinates: 22, −52, 54) (T = −2.55, P_FDR_= 0.05). In **Figure 5**, the magnitudes of functional connectivity between each pair of ROIs and significant clusters are shown.

**Figure 5.**
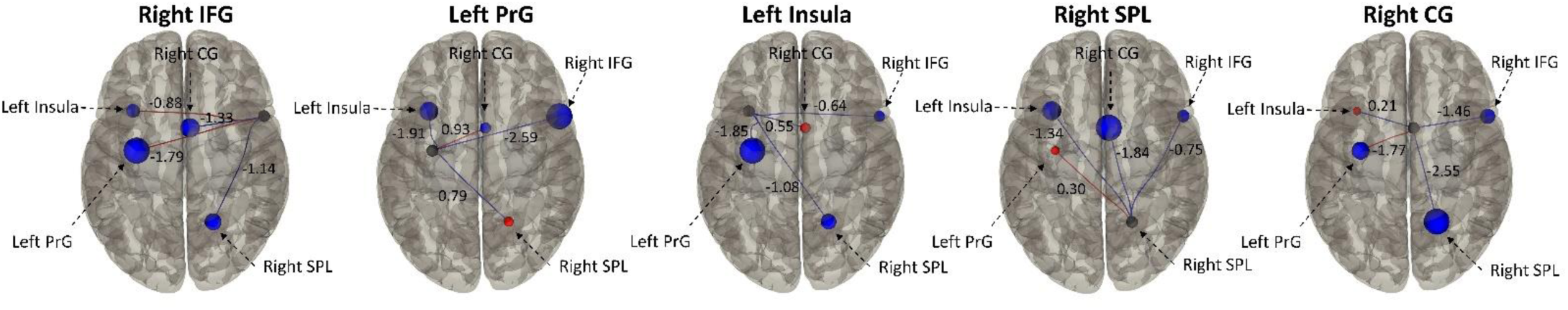
Cue reactivity network between areas with subjective-neural associations. PrG-IFG and CG-SPL showed significantly declined functional connectivity during the food cue reactivity task (thresholded at P_FDR_ < 0.05). ***Note.*** *PrG* precentral gyrus, *IFG* inferior frontal gyrus, *SPL* superior parietal lobule, *CG* cingulate gyrus.

### 3.4. Group Factor Analysis

We conducted a group factor analysis (GFA) to explore whether a broader coherent pattern could be identified for the association between psychological/behavioral evaluations, demographic features, and neural correlates of RFC among overweight/obese participants when exposed to food cues. Five latent constructs (factors) were extracted using GFA **(Figure 6-A)**. Factors 1, 2, 3, and 5 comprised neural variables that represented regional activities within regions showing a positive correlation with RFC scores. These factors accounted for 29.1% of the total variance. Factors 2 and 4 contained psychological variables, together explaining 12.23% of the variance across measures. Intriguingly, GFA unveiled a cross-unit latent factor that accounted for 9.3% of the variance across neural and psychological measures (factor 2) **(Figure 6-A)**.

**Figure 6.**
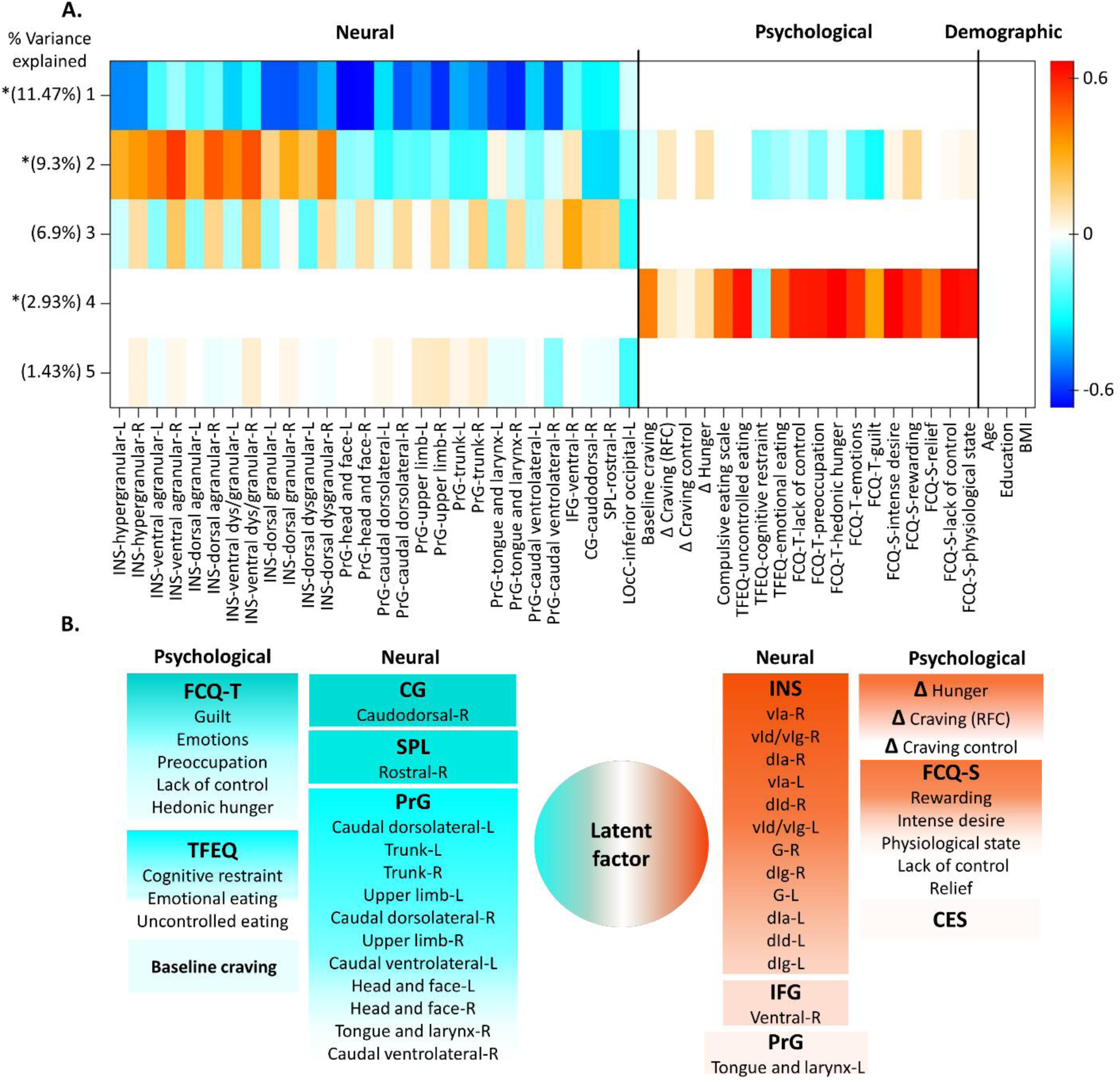
Visualization of factor loadings derived from the Group Factor Analysis (GFA) results. **(A)** The loadings of three group variables across five GFA factors are shown. **(B)** Loadings of neural and psychological variables on the cross-unit latent factor (Factor 2), showing the interplay between neural and psychological datasets. Positive loadings (shades of red) and negative loadings (shades of blue) indicate variables contributing to and detracting from the latent factor, respectively. Relevant associations included positive loadings of the insula and FCQ-S (particularly the rewarding subscale) in contrast to the negative loadings of PrG and FCQ-T (especially the guilt subscale). ***Note.*** *GFA* group factor analysis, *L* left and *R* right, *INS* insular gyrus, *INS-G* hypergranular insula, *INS-vIa* ventral agranular insula, *INS-dIa* dorsal agranular insula, *INS-vId/vIg* ventral dysgranular and granular insula, *dIg* dorsal granular insula, *dId* dorsal dysgranular insula, *PrG* precentral gyrus, *IFG* inferior frontal gyrus, *PCC* posterior cingulate cortex, *SPL* superior parietal lobule, *LOcC* lateral occipital cortex, *RFC* responsiveness to food cues, *CES* compulsive eating scale, *TEFQ* three-factor eating questionnaire, *FCQ-T* food craving questionnaire-trait, *FCQ-S* food craving questionnaire-state, *BMI* body mass index.

All subregions of the insula had a robust positive loading on this latent factor. Additionally, the subscales of the food craving questionnaire-state (FCQ-S), with a notable loading on the ‘anticipation of reward’ subscale, RFC scores, changes in hunger level, and IFG activity showed a somewhat less prominent, but still positive loading on this factor. Conversely, the caudodorsal cingulate, SPL, and subregions of the PrG, excluding the left tongue and larynx area, had negative loadings on this factor, followed by all subscales of the food craving questionnaire-trait (FCQ-T), particularly the ‘guilt’ subscale, cognitive strategies aimed at restricting food intake (a subscale of the three-factor eating questionnaire, TFEQ) and baseline craving. **Figure 6-B** illustrates the interplay between neural and psychological variables with respect to the latent factor (factor 2) (see **Supplementary Figure 3** for heat maps).

## 4. Discussion

With the primary aim of a deeper understanding of food cue-dependent interoception processing encoded in the insula, this study delved into the complexities of this brain region and its interplay with both neural and psychological/behavioral components among overweight and obese participants.

Our analysis unveiled significant associations between psychological and brain activity features, providing insights into the neural and psychological underpinnings of interoceptive processing modulated by a food cue reactivity task.

The primary findings of the whole brain analysis revealed significant activations in several brain regions, including left medioventral fusiform gyrus (FuG), left medial amygdala, right posterior cingulate cortex (PCC), and left dorsolateral area of superior frontal gyrus (SFG) in response to food cues compared to neutral cues, largely consistent with a large body of prior studies, illustrating that food-related cues engage a network of brain regions involved in reward/emotion/motivation (Bruce et al., 2010; Devoto et al., 2018; Dimitropoulos, Tkach, Ho, & Kennedy, 2012; Isabel García-García et al., 2014; Pursey et al., 2014; Rothemund et al., 2007; Stoeckel et al., 2008) and visual processing (Baboumian et al., 2019; Connor, Egeth, & Yantis, 2004; Huerta, Sarkar, Duong, Laird, & Fox, 2014). In the following, we discuss the activation of these regions in response to food cues.

Increased activity of the FuG may indicate modulation of neural circuits associated with visual processing of food cues, possibly reflecting salience attribution given to foods and evaluation of the rewarding effect of stimuli in our study sample (Baboumian et al., 2019; Connor et al., 2004; Drewnowski, Brunzell, Sande, Iverius, & Greenwood, 1985; Higgs, Rutters, Thomas, Naish, & Humphreys, 2012; Siep et al., 2009), which is consistent with a substantial increase of the subjective experience of craving and hunger levels following food cue exposure (Aviram-Friedman, Kafri, Baz, Alyagon, & Zangen, 2020).

Furthermore, a larger activity of the PCC associated with personally salient cues has been shown in previous neuroimaging studies (Pearson, Heilbronner, Barack, Hayden, & Platt, 2011; Sadler et al., 2023), possibly reflecting enhanced attention to internal body states (Tregellas et al., 2011) and emotional/motivational processing (Litt, Plassmann, Shiv, & Rangel, 2011; Maddock, Garrett, & Buonocore, 2003).

Furthermore, enhanced activation of the dorsolateral area of the SFG was observed. Previous studies have indicated diminished activation of the prefrontal control network in response to food cues in overweight/obese individuals (Brooks, Cedernaes, & Schiöth, 2013; Christensen, Harding, Voigt, Chong, & Verdejo-Garcia, 2022; I García-García et al., 2013; Song et al., 2022; Veit et al., 2021), it is also evident that in a satiation state, these tend to show enhanced activation in more anterior regions of the prefrontal cortex compared to their normal weight counterparts (Del Parigi et al., 2002; Devoto et al., 2018; Pursey et al., 2014), potentially indicating a cognitive readiness for food consumption (Goldschmidt et al., 2018; Nederkoorn & Jansen, 2002), which does not necessarily contribute to enhanced cognitive control over excessive eating tendencies. This is evident by the reduced self-reported craving control scores we observed, indicating that exposure to food cues led our volunteers to perceive a diminished ability to regulate food-related desires. This observation is also consistent with previous studies that emphasized the impact of food cue exposure on subsequent food intake, independent of factors such as BMI, dietary restraint (Boswell & Kober, 2016), and current hunger levels (Nederkoorn & Jansen, 2002).

Furthermore, enhanced activation of the amygdala, an essential part of the reward circuitry, suggests an emotional engagement with the rewarding value of food cues (Baxter & Murray, 2002; Killgore et al., 2003; Mahler & Berridge, 2012; Richter-Levin & Akirav, 2003; Siep et al., 2009). Moreover, the amygdala, along with the insula, as the main region of the saliency network (SN), is relevant for the integration of sensory inputs to guide adaptive appropriate behavior in response to emotionally salient stimuli (Janak & Tye, 2015). However, our primary group-level analysis did not reveal insula activation following the presentation of food cues. This might be explained by the fact that the participants in our study were instructed to abstain from eating and maintain a neutral hunger level for 1-2 hours before scanning to standardize pre-scan conditions (Smeets et al., 2019), ensuring they were in a state of either absent or negligible hunger. This instruction aimed to mitigate the impact of potential variabilities associated with different states of hunger that could influence results, as observed in previous research (Demos, Heatherton, & Kelley, 2012; Gautier et al., 2000). Previous studies have emphasized the involvement of the insula in conscious craving (Siep et al., 2009), along with its association with the motivational value of food and further food intake (Small, Zatorre, Dagher, Evans, & Jones-Gotman, 2001; Tang et al., 2012), particularly in the fasted state (Del Parigi et al., 2002). Therefore, the insignificant cue-provoked insula response likely indicates a relatively modest rewarding value of food cues due to participants’ satiation states. Nevertheless, we observed higher insula activation in individuals with a heightened subjective response to food cues, which is consistent with previous studies by indicating that overweight/obese participants who exhibited increased responsiveness to food cues (RFC) also demonstrate higher activation of the insula, suggesting heightened motivation to consume food (Johnson, 2013).

Further, to specifically explore neural correlates of the RFC ratings, reflecting visceral interoceptive processing, we employed voxel-wise whole brain correlation analysis. This approach revealed that overweight/obese individuals with higher RFC show enhanced regional brain activities within the precentral gyrus (PrG), insula, inferior frontal gyrus (IFG), caudodorsal cingulate, and superior parietal lobule (SPL).

While multiple regions have shown a significant association with RFC measures in response to food cues, the insular cortex has long been considered a core region within the interoceptive network due to its prominent role in perceiving and integrating body signals for the maintenance of metabolic homeostasis (Craig, 2002). Altered insula activation has been frequently observed in cued task studies and is hypothesized to directly contribute to disruptions in weight control and obesity (Avery et al., 2015; Simmons, Martin, & Barsalou, 2005).

In addition to the insula, guided by previous frameworks (Barrett & Simmons, 2015; Showers & Lauer, 1961) highlighting the tight link between insular interoceptive processing and generation of motor actions (Barrett & Simmons, 2015; Singer, Critchley, & Preuschoff, 2009), and based on our findings indicating strong correlations of the insula and PrG with RFC scores, we further examined the blood oxygenation level-dependent (BOLD) signal within all subregions of the PrG − a region primarily responsible for motor processing, planning, and executing voluntary movements − in response to food cue exposure, suggesting a potential involvement in establishing motor readiness for ingestion (Kahathuduwa, Boyd, Davis, O’Boyle, & Binks, 2016; Karhunen, Lappalainen, Tammela, Turpeinen, & Uusitupa, 1997; McCaffery et al., 2009; Sadler et al., 2023; G.-J. Wang, Volkow, & Fowler, 2002). Specifically, the association between disinhibited eating and heightened BOLD activity in the motor cortices has previously been demonstrated in neuroimaging studies (Aviram-Friedman, Astbury, Ochner, Contento, & Geliebter, 2018; Zhao et al., 2017), especially in the context of highly palatable foods (Stice et al., 2010). The subsequent discussion predominantly centers around insula and PrG, along with their respective subregions, as our ROIs.

For insular subregions, an increased BOLD response was detected in the agranular anterior regions in participants with higher RFC scores. This aligns with the theoretical framework about the importance of the anterior insula in visceral interoceptive awareness, as postulated by Craig (2003), and supported by recent functional imaging studies (Huang et al., 2021). Moreover, the dorsal agranular insula showed the most robust correlation with RFC ratings, succeeded by the dorsal dysgranular, and subsequently ventral granular regions, constituting the dorsal anterior segment of the insula. These findings tentatively imply a hierarchical processing mechanism within the insula, where different subregions contribute to the modulation of food cue reactivity based on the level of interoceptive integration. This is consistent with the proposed functional organization model of the insula which posits a sequential flow of interoceptive information processing from the granular to the dysgranular and ultimately to the agranular portions of the insula (Craig, 2009).

The high correlation between RFC and the agranular insula can be interpreted within the framework of the embodied predictive interoception coding (EPIC) model (Barrett & Simmons, 2015). Based on this model, in the normal functioning brain, the anterior insula seeks to minimize the disparity between pre-existing predictions (shaped by past experiences) and actual sensations (incoming sensory inputs), known as prediction error. This is accomplished through the anterior insula due to its involvement in the intrinsic executive control and attention networks, facilitated by its rich connections with the prefrontal cortex (Seeley et al., 2007; Sridharan et al., 2008; Uddin et al., 2014), and involves redirection of the focus of attention, adjustment of internal predictions, or active induction of the predicted sensations through self-generated motor actions (Barret and Simmons, 2015). Further, this model offers insights into how the intensified activity of the anterior insula, by aiming to minimize the prediction error, ultimately leads to disturbed behaviors. Thus, the current finding in conjunction with increased activation of the PrG, along with the substantial increase of RFC and reduced self-reported craving control, provides compelling supportive evidence for central assumptions of this model in the food cue reactivity domain.

For specific PrG subregions, the ROI analysis revealed substantial positive associations between a greater hemodynamic response in PrG subregions (including head and face, upper limb, caudal dorsolateral, and tongue and larynx areas) and RFC scores. Rapuano et al. (2016) concluded that increased activity of oral somatosensory and motor cortices in obese adolescents in response to food commercials may contribute to motor preparation for food consumption (Rapuano et al., 2016). Interestingly, the authors argue that the peak activations observed in bilateral sensorimotor regions have been previously documented in fMRI studies investigating neural correlates of oral sensorimotor tasks such as lip, tongue, larynx, and jaw movements (Grabski et al., 2012), or mastication (Takahashi, Miyamoto, Terao, & Yokoyama, 2007). Here, we deepened these findings by providing further evidence of similar activity in response to static food-related cues in overweight/obese individuals.

It is essential to note that, as a major node of the SN, the anterior insula coordinates large-scale brain networks and has a causal role in mediating the transition between the default mode network (DMN) and the central executive network (CEN) to optimize executive control (Margulies & Uddin, 2019; Molnar-Szakacs & Uddin, 2022; Uddin, 2015). By receiving interoception signals, salient external stimuli, as well as emotional and cognitive signals from various cortical and subcortical regions (Craig, 2002), the insula enables the initiation of switching between the CEN and DMN (Uddin, 2015). To effectively deal with stimulus-driven tasks, Individuals need to adjust the allocation of cognitive resources to inhibit default responses (deactivating the DMN), and establish new coping behaviors (activating the CEN) aimed at generating appropriate responses toward relevant stimuli (Sridharan et al., 2008; Uddin, 2015). Nevertheless, overweight individuals often show an approach-avoidance pattern of attention allocation to food-related cues, leading to impaired inhibitory control and functional switching between DMN and CEN, thereby resulting in habitual eating or tendencies toward overeating (Werthmann et al., 2011).

Taken together, and considering other regions showing positive neural-subjective associations, we postulate that the findings of a stronger BOLD signal in the anterior insula (a key region of the SN) (Uddin, 2015), along with simultaneous higher activation in the caudodorsal cingulate (a key node of the DMN) (Yu et al., 2011), IFG (a central component of the CEN) (Chambers, Garavan, & Bellgrove, 2009), as well as the PrG, which involves motor regions associated with voluntary movement and motor preparation (Saito et al., 2019), and the SPL, which is involved in visually guided motor actions (Hahn, Ross, & Stein, 2006), collectively may point towards a potential failure of the anterior insula in modulating interference and facilitating functional switching between the DMN and CEN in response to food stimuli, which is essential for guiding adaptive behavior, a conclusion supported by recent studies (Huang et al., 2021; Sridharan et al., 2008; Trautwein, Singer, & Kanske, 2016).

This interpretation of the findings would be strengthened by incorporating results from seed-to-seed analyses. The observed weakened functional connectivity between the caudal dorsolateral PrG and the ventral IFG suggests compromised inhibitory control in overweight/obese individuals when exposed to food cues, potentially enhancing their vulnerability to impulsive eating behaviors (Lawrence et al., 2012; Reyes, Peirano, Peigneux, Lozoff, & Algarin, 2015; Veit et al., 2021). The involvement of the IFG in voluntary motor control, as indicated by its role in timing the initiation of motor tasks, has been previously investigated (Rao et al., 1993). In this connection, women with food addiction show decreased connectivity in the sensorimotor network, particularly in regions associated with inhibitory control, such as the IFG (Hampshire, Chamberlain, Monti, Duncan, & Owen, 2010).

Similarly, we observed diminished connectivity between the caudodorsal cingulate and the rostral SPL. There is evidence for widespread connections between the PCC and the prefrontal cortex (Leech, Kamourieh, Beckmann, & Sharp, 2011; Summerfield, Hassabis, & Maguire, 2009), as well as frontoparietal regions involved in the executive control network (Vincent, Kahn, Snyder, Raichle, & Buckner, 2008), suggesting a cognitively active role of the PCC in adjusting and controlling behaviors (Leech, Braga, & Sharp, 2012; Pearson et al., 2011; Raichle & Snyder, 2007). Conversely, the SPL has been linked with the integration of somatosensory and motor information, especially concerning visually guided motor actions (Glover, 2003; Hahn et al., 2006), as well as visual attention toward salient stimuli (Behrmann, Geng, & Shomstein, 2004). This might suggest an enhanced preoccupation with food cues, observed in overweight/obese individuals, consequently raising the motivational rewarding value of food cues (Hardman et al., 2021). Previous studies indicated that this intensified focus on salient stimuli may suppress cognitive regions within the DMN, leading to declined inhibitory control and increased impulsive decision-making in response to such stimuli (McFadden, Tregellas, Shott, & Frank, 2014).

Lastly, the group factor analysis (GFA) identified a specific cross-unit latent factor across neural and psychological/behavioral measures that significantly influence eating tendencies toward food consumption. This latent factor is shaped by a combination of neural and psychological/behavioral variables, indicating the complexity of behaviors related to food consumption. The involved variables can be classified into two distinct groups, acting as ‘suppressors’ and ‘facilitators’ in shaping eating tendencies towards food consumption. Suppressor variables encompass both neural (activation of the PrG, SPL, and caudodorsal cingulate) and psychological measures (FCQ-T, cognitive restraint of eating in TFEQ, and baseline craving), suggesting that an increase of these variables contributes negatively to the factor identified by GFA (factor 2). Conversely, higher levels of facilitator variables, including both neural (activation of the insula and IFG) and psychological measures (FCQ-S, RFC, and cue-induced hunger change), prompt this factor.

Notably, scores of FCQ-T and FCQ-S exerted contrasting impacts on the latent factor (factor 2). While FCQ-T exhibited negative loadings, FCQ-S demonstrated positive loadings on the factor, suggesting that individuals with high trait and state cravings may have different neuropsychological responses to food cue exposure. This may imply that factors suppressing overeating tendencies in connection with food-related cues might be particularly prominent in high-trait cravers — individuals who experience trait-like features associated with more frequent and intense cravings (Innamorati et al., 2014) and who are also more inclined to employ restrictive eating strategies (Goldschmidt et al., 2018; Nederkoorn & Jansen, 2002). In contrast, overweight/obese individuals with high-state craving — reflecting transient changes in the physiological state following food deprivation and exposure to food cues (Meule, Lutz, Vögele, & Kübler, 2014) — may have impaired inhibitory control mechanisms and excessive reward-related eating, rendering them more vulnerable to excessive weight gain (Mela, 2001; Meule et al., 2018; Rodin & Slochower, 1976). Indeed, high-trait cravers are more likely to be less responsive to food cues, as they may exert more effort in regulating eating tendencies, compared to their counterparts with enhanced high-state craving. This may be attributed to their familiarity with cognitive strategies aimed at restricting food intake, potentially signifying a cognitive readiness for possible food consumption.

In conclusion, consistent with previous studies, our results add further evidence to the view that pathological interoceptive processing in overweight/obese individuals, associated with viscerosensory consequences such as subjective sensations of hunger and craving, may be attributed to insular dysfunction, resulting in action tendencies towards food consumption, probably due to the hyperactivation of motor-related regions rather than homeostatic regulation mechanisms. Specifically, this dysfunction may be attributed to the impaired function of the anterior insula, as a main region of the interoceptive processing network, to control action tendencies toward food consumption.

Our novel finding indicates that participants with higher FCQ-T scores may have a different neuropsychological profile than those with higher FCQ-S scores, potentially influencing how individuals with distinct psychological and neural profiles perceive and respond to food-related stimuli. Identification of these differences is essential for informing interventions aimed at promoting healthier eating behaviors and exerting control over overeating tendencies.

Future research might utilize these findings to conduct a more precise examination of interoceptive processing and identify additional factors that mediate the distinctive patterns of regional brain responses to food-related stimuli in order to develop more targeted interventions for managing overeating and obesity, considering both facilitatory and suppressor factors.

However, our study had some limitations, including the absence of a control group with lower BMI for direct comparisons, and multiple exploratory analyses in a moderately sized sample population. Caution should be taken in generalizing the results, and further studies with well-matched control groups and also confirmatory analyses focused on the regions and networks identified in this study are needed for a better understanding of the neurobiological mechanisms underlying interoceptive processing in the context of obesity. Additionally, future studies with larger samples and a broader examination of brain networks involved in interoception would provide a more comprehensive insight into respective mechanisms.

## Ethics statement

The current study was approved by the ethics committee of research, Iran University of Medical Sciences (IR.IUMS.REC.1396.0459). All participants provided written informed consent to participate in this study.

## Acknowledgment

The authors would like to express their gratitude to all research assistants and participants for their collaboration in the study. A special thanks to Hosna Tavakoli, Peyman Ghobadi-Azbari, and Mohsen Ebrahimi for their valuable contributions throughout the project.

## Author Contributions

All authors contributed to the conceptualization of the research. NM conducted the research and collected the data with support from the Iranian National Brain Mapping Laboratory. The manuscript was written by NM, and HE was the principal supervisor for editing the manuscript. MN, SR, and AF reviewed and edited the paper and all authors read and approved the submitted manuscript.

## Data availability statement

The data supporting the findings of this study will be available from the corresponding authors by request from any qualified investigator.

## Declaration of generative AI and AI-assisted technologies in the writing process

During the preparation of this work, the author(s) used ChatGPT developed by OpenAI to refine content and ensure English grammar correctness. After using this tool, the author(s) reviewed and edited the content as needed and took full responsibility for the content of the publication.

